# Genome sequencing in the Parkinson’s disease clinic

**DOI:** 10.1101/2021.11.23.21266755

**Authors:** Emily J. Hill, Laurie A. Robak, Rami Al-Ouran, Jennifer Deger, Jamie C. Fong, Paul Jerrod Vandeventer, Emily Schulman, Sindhu Rao, Hiba Saade, Rainer von Coelln, Harshavardhan Doddapaneni, Sejal Salvi, Shannon Dugan-Perez, Donna M. Muzny, Amy McGuire, Zhandong Liu, Richard Gibbs, Chad Shaw, Joseph Jankovic, Lisa M. Shulman, Joshua M. Shulman

## Abstract

**Background and Objectives:** Genetic variants impact both Parkinson’s disease (PD) risk and manifestations. While genetic information is of potential interest to patients and clinicians, genetic testing is rarely performed during routine PD clinical care. The goal of this study was to perform genome sequencing and examine patient interest in comprehensive genetic testing for PD in 2 academic movement disorder clinics.

**Methods:** In 208 subjects with PD (age=63 years, 67% male), genome sequencing was performed and filtered using a custom panel, including 49 genes associated with PD, parkinsonism, or related disorders, as well as a 90-variant PD genetic risk score. Separately, 231 patients (age=67 years, 63% male) were surveyed on interest in genetic testing at baseline and in response to vignettes covering (i) familial risk of PD (*LRRK2*); (ii) risk of PD dementia (*GBA*); (iii) PD genetic risk score; and (iv) secondary, medically-actionable variants (*BRCA1*).

**Results:** Genome sequencing revealed a *LRRK2* variant in 3.4% and a *GBA* risk variant in 10.1% of our clinical sample. The genetic risk score was normally distributed, identifying 42 subjects with high risk of PD. Medically-actionable findings were discovered in 2 subjects (1%). In our survey, the majority (82%) responded they would share a *LRRK2* variant with relatives. Most registered unchanged or increased interest in testing when confronted with potential risk for dementia or medically- actionable findings, and most (75%) expressed interest in learning their PD genetic risk score.

**Discussion:** Our results highlight broad interest in comprehensive genetic testing among patients with PD and may facilitate integration of genome sequencing in clinical practice.

## Introduction

Advances in genetic knowledge have implicated more than 100 genes in Parkinson’s disease (PD) risk, including both rare and common variants.^1,2^ Along with potential implications for counseling families, identification of these genetic risk factors may also impact clinical decision-making. Several clinical trials for targeted therapies in PD are now recruiting patients with *GBA-* PD or *LRRK2-*PD, motivating large-scale efforts to identify eligible subjects.^3^ In addition, numerous studies highlight links between specific genetic variants and heterogeneous PD symptoms. For example, *GBA*-PD is characterized by more rapid disease progression and an increased risk of dementia.^4–7^*APOE* genotype, which has an established role in clinical risk stratification for Alzheimer’s disease^8^, is also associated with Lewy body dementia.^9,10^ In *LRRK2*-PD, several studies have suggested a more benign disease course, while others reveal a higher likelihood of the postural instability gait difficulty motor phenotype.^11,12^ Lastly, while most common PD risk variants (i.e., population allele frequency > 1%) have only modest effect sizes in isolation, a genetic risk score incorporating dozens of single nucleotide polymorphisms (SNPs) predicts accelerated PD progression^13,14^ and may also facilitate early diagnosis.^15^ Because it is derived from common variant genotypes, the PD genetic risk score may be applicable to many patients with PD. Similar algorithms have been clinically validated for risk assessment in other common and complex genetic disorders, such as cardiovascular disease.^16,17^

Most current commercial gene panels for PD are largely restricted to testing for monogenic causes of Mendelian PD. The available tests also vary widely in the number of genes and variants tested, frequently include loci with uncertain evidence, and do not include common polymorphisms or assess a genetic risk score.^18^ Compared with targeted panel assays, genome sequencing comprehensively examines both rare and common variants in most genes. In addition, results can be readily filtered and reported based on all available evidence, making possible iterative analytic updates following new discoveries and without requiring an additional blood draw or new data generation. The American College of Medical Genetics and Genomics (ACMG) recommends offering testing for genes conferring risk of a serious medical condition for which there is an effective intervention or need for surveillance.^19^ Such “medically-actionable” genetic findings unrelated to PD can be readily screened using genome sequencing.

Despite the growing potential to impact clinical decision making, genetic testing is rarely employed in the routine clinical assessment of PD^20^, yet most patients with PD have a strong desire to know their genetic information.^21–23^ Besides potential barriers for reimbursement, many neurologists feel poorly prepared to provide counseling to patients regarding results.^24^ In addition, other issues that may arise from comprehensive testing using genome sequencing have not been fully explored, such as disclosure of genetic risk scores or potential discovery of medically actionable findings unrelated to PD diagnosis. In order to address these gaps, it is essential to document not only the spectrum of genetic findings that would be expected in clinical practice, but also the possible reactions of patients to such results.

## Methods

### Subject Recruitment

Subjects were recruited from 2 academic movement disorder centers (Baylor College of Medicine and University of Maryland School of Medicine). During routine clinic visits between 2013-2019, patients with PD who were diagnosed by a movement disorder specialist were offered participation in a DNA bank repository. Demographics and selected clinical details were collected through chart review and confirmed based on an unstructured interview at the time of sample collection in the clinic, including family history of PD (defined as any affected blood relative), age at symptom onset, and results from cognitive screening. The DNA bank repository protocol permits genome sequencing for research, but does not allow for disclosure of results. Therefore, in a separate study conducted between 2018-2019, PD patients from both clinics were recruited for an approximately 15-minute survey in which responses were recorded to several clinical vignettes including genetic testing results (below). Individuals with dementia and those who had previously undergone clinical genetic testing for PD were excluded from the survey study. There were 231 subjects who contributed DNA samples for genome sequencing and 208 who completed the survey. Twenty-eight subjects participated in both parts of the study.

### Standard Protocol Approvals, Registrations, and Patient Consents

All subjects provided informed consent. The Institutional Review Boards at Baylor College of Medicine and University of Maryland approved both the biospecimen collection / sequencing and survey study included in this report.

### Custom PD Gene Panel

We developed a custom PD gene panel for filtering sequencing results and creation of personalized PD genome risk profile reports (Supplemental Table 1). We included 5 categories of genes/variants, including both (1) 5 highly-penetrant PD risk genes (*SNCA, LRRK2, VPS35, PARK2/PRKN, PINK1*) and (2) moderately-penetrant risk genes and PD modifiers (*GBA, SMPD1, APOE*). We included *APOE* in this latter category as it has an established association with cognitive impairment in PD.^9,25^ Because idiopathic PD clinical manifestations sometimes overlap with other genetic syndromes that cause parkinsonism (e.g., dopamine-responsive dystonias^26^), we also included an additional category (3) including 41 such genes (Supplemental Table 1). All genes selected for the panel were based on careful literature review with the following inclusion criteria: at least one pathogenic variant reported in 3 separate families by 2 or more independent groups or at least 1 large sequencing study revealing a significant association with PD or parkinsonism. For the PD genetic risk score (category 4), we considered 90 risk variants identified by PD genome-wide association study meta-analysis (GWAS), weighted by their individual odd ratios for PD.^16,27^ We divided the genetic risk scores into low, medium, and high risk based on quintiles (1^st^ quintile defined as low risk, 2^nd^-4^th^ quintile as medium risk, and 5^th^ quintile as high risk)^28^. Subject groups with a high versus low genetic risk score were compared in terms of age at onset and Montreal Cognitive Assessment (MoCA) score using the student’s t-test. Lastly (category 5), our panel included 59 genes unrelated to PD, but which were recommended for secondary reporting by the ACMG at the time of this analysis.^29^

### Genome Sequencing

DNA was extracted from peripheral blood lymphocytes or saliva samples using standard protocols. Whole genome sequencing data was generated for 220 samples at BCM-HGSC using established library preparation and sequencing methods. Libraries were prepared using KAPA Hyper PCR-free library reagents (KK8505, KAPA Biosystems Inc.) on Beckman robotic workstations (Biomek FX and FXp models). Briefly, DNA (750 ng) was sheared into fragments of approximately 200-600 bp using the Covaris E220 system (96 well format, Covaris, Inc. Woburn, MA) followed by purification of the fragmented DNA using AMPure XP beads. A double size selection step was employed, with different ratios of AMPure XP beads, to select a narrow size band of sheared DNA molecules for library preparation. DNA end-repair and 3’-adenylation were then performed in the same reaction followed by ligation of the Illumina unique dual barcodes adapters (Cat# 20022370) to create PCR-Free libraries, and the library was run on the Fragment Analyzer (Advanced Analytical Technologies, Inc., Ames, Iowa) to assess library size and presence of remaining adapter dimers. This was followed by qPCR assay using KAPA Library Quantification Kit (KK4835) using their SYBR® FAST qPCR Master Mix to estimate the size and quantification. WGS libraries were sequenced on the Illumina NovaSeq instrument utilizing the S4 flow cells with 24 libraries in each lane. Sequencing reagents used included the NovaSeq 6000 S4 Reagent Kit (Cat#20012866) and NovaSeq Xp 4-Lane Kit (Cat# 20021665). Libraries were loaded at an average concentration of 365 pM to generate 150 bp paired-end reads. Sequence data was processed using HgV (‘Mercury V’, version.17.5), the HGSC workflow management system, and mapped to human reference build hg19.^30,31^ Unique aligned sequences in these samples varied between 69.6 Gb – 285.7 Gb, with an average of 129.89Gb per sample. The average median insert size for these samples was 420 bp and the average mode insert size was 399 bp.

### Variant Filtering/Analysis

All variants, including both exonic or intronic variants, were initially annotated using ClinVar (http://www.ncbi.nlm.nih.gov/clinvar/) and Human Gene Mutation Database (HGMD) in April 2019, supplemented by literature review and discussion by our multi-disciplinary team, which included a clinical geneticist, a clinical genetic counselor, and basic scientists.^32^ With the exception of *GBA* (see below), all variants considered as pathogenic in this study were annotated in ClinVar as either “pathogenic” / “likely pathogenic” or in HGMD as “DM”, indicating disease causing. In the case of *GBA*, database annotation is largely based on risk for Gaucher disease; however, we additionally considered 2 *GBA* variants, E365K and T408M, with strong literature support for increased risk for PD but which are non-pathogenic for Gaucher.^33,34^ For all variants, we additionally performed literature review to document consistent reports from multiple independent families and/or large cohort sequencing studies, as well as cases where variants had support from experimental validation in model organisms. This supportive evidence is documented in Supplemental Table 2. Following exclusion of ClinVar “benign” or “likely benign” variants, all other identified findings were considered as variants of uncertain significance (VUS) and are reported in Supplemental Table 3.

### Survey Study

The survey was comprised of 4 clinical vignettes concerning genetic testing (See Supplement Information for complete survey). The vignettes highlighted the following scenarios: family risk of PD due to a *LRRK2* variant (vignette 1); personal dementia risk in a carrier of a *GBA* variant (vignette 2); negative findings for highly-penetrant variants and reporting of the genetic risk score (vignette 3); and discovery of a medically-actionable variant in *BRCA1* (vignette 4). These vignettes were designed both as an educational tool and to assess subject reactions, including how patient interest in genetic testing may be impacted by different results. Vignettes and follow-up questions were read aloud to subjects. Interest-level for comprehensive genetic testing was assessed based on a Likert scale (0, “much less interested” to 5, “much more interested”). Qualitatively, we also recorded comments from subjects on factors influencing their decision-making. Comments responding to at least 1 vignette were recorded from 133 out of 231 subjects. At the end of the survey, we requested feedback on how doctors might better prepare patients for genetic testing in PD.

### Data Availability

Complete results from the survey are available in Supplemental Table 4. For all subjects who consented for public data sharing, sequencing data are in process for release in relevant genomic databases. The complete sequencing data are also available on request by contacting the corresponding author, Dr. Shulman.

## Results

### Genome Sequencing

We performed genome sequencing in 208 unselected, unrelated subjects with PD. Subject demographics and clinical characteristics are summarized in Table 1. For filtering and annotation of personal genomes, we developed a custom PD gene and variant panel including 5 categories of reportable genetic risk (Methods). The gene panel was designed to facilitate generation of individual PD Genome Sequencing Reports (Figure 1). Since genome sequencing was conducted under a protocol that did not allow for disclosure of results, the individual findings instead informed our development of several clinical vignettes that we subsequently presented to a largely non-overlapping sample of subjects with PD.

**Table 1.** Subject Characteristics.

|  | Sequencing | Survey |
| --- | --- | --- |
| Subjects, n | 208 | 231 |
| Age, mean (SD) | 63 (9) | 67(9.9) |
| Sex, % male | 67% | 63% |
| Disease duration in years, mean (SD) | 7 (5) | 9 (5) |
| Race, % European Ancestry | 90% | 63% |
| Family history of PD<sup>1</sup>, % | 27% | 24% |

<sup>1</sup>Family history was defined as any blood relative with Parkinson's disease.

**Figure 1.**
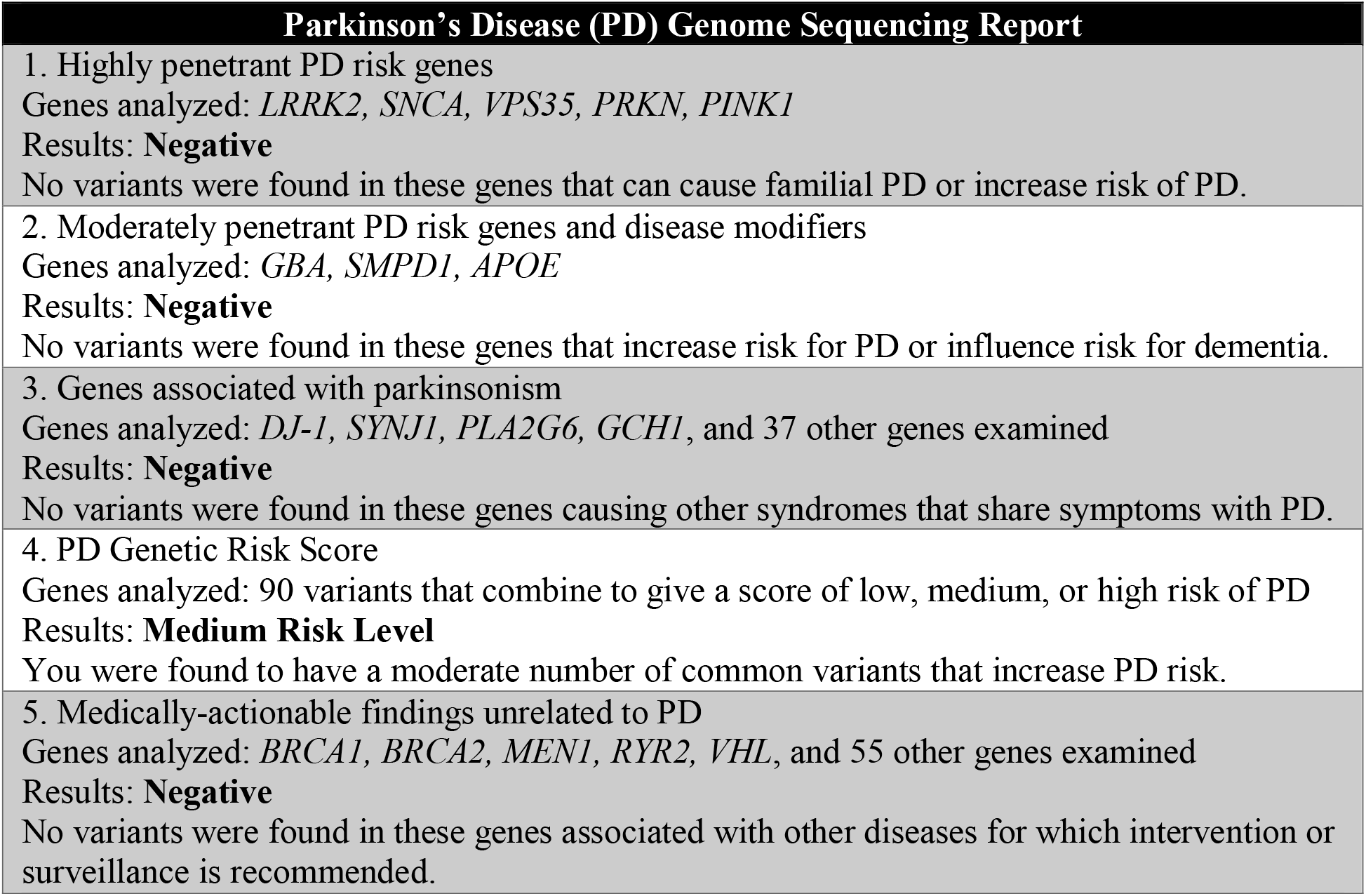
Sample PD genome sequencing report. This report represents the most common results based on application of our customized PD gene panel (See also Supplemental Table 1 for all genes included in categories 1-3).

Among highly penetrant gene variants (category 1; see also vignette 1, below), our sample included 7 subjects with a *LRRK2* pathogenic variant (3.4%) as well as 1 heterozygous carrier of a pathogenic *PRKN* variant. An additional 21 subjects (10.1%) were discovered to have a VUS in at least one category 1 gene (Supplemental Table 3). We next examined variants in genes associated with moderate risk of PD as well as disease modifiers (category 2; vignette 2). We discovered 21 carriers of *GBA* pathogenic variants, and 2 subjects with an *SMPD1* variant. Notably, one subject was a carrier of both a *GBA*^*E356K*^ and *LRRK2*^*M1646T*^ variant. For *APOE*, 47 subjects were heterozygous for the *e4* allele. Two individuals were homozygous for *APOE-e4*, but based on available clinical documentation, both individuals were not known to have cognitive impairment. Table 2 shows all reportable pathogenic variants in genes from categories 1 and 2. In Category 3, we examined other genetic causes of parkinsonism; however, no individuals in our sample were carriers of such variants.

**Table 2.**
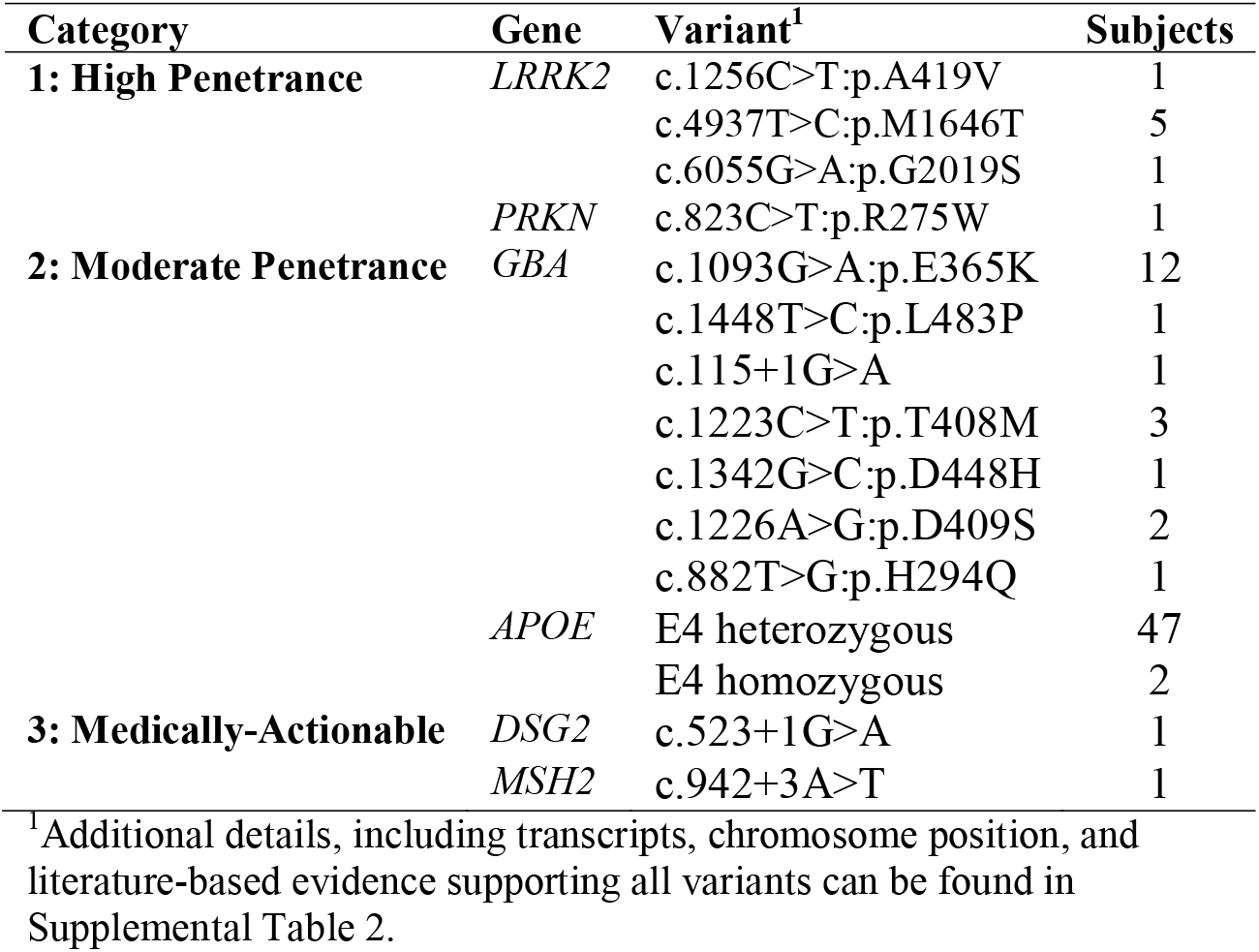
Sequencing Results.

We next examined the 90-variant PD genetic risk score (category 4), which was normally distributed in our sample (Figure 2). After dividing into quintiles^28^, there were 42 subjects each with a high or low genetic risk score. Of note, we found no significant difference between age at symptom onset or level of cognitive function in comparisons of the low-versus high-risk groups (p=0.6 and p=0.2, respectively), despite reports of such associations in much larger sample sizes.^35,36^ As expected, most subjects (n=124) had a score consistent with moderate risk, and in for most of these (n=109), no other genes were flagged from categories 1-3 (representative report, Figure 1; vignette 3). Lastly, we examined 59 ACMG medically-actionable findings, unrelated to PD risk (category 5; vignette 4). We discovered 2 individuals harboring such alleles, including a *DSG2* variant associated with arrhythmogenic right ventricular cardiomyopathy as well as a variant in *MSH2* which causes Lynch syndrome, conferring an increased risk for colon and other cancers.^29^

**Figure 2.**
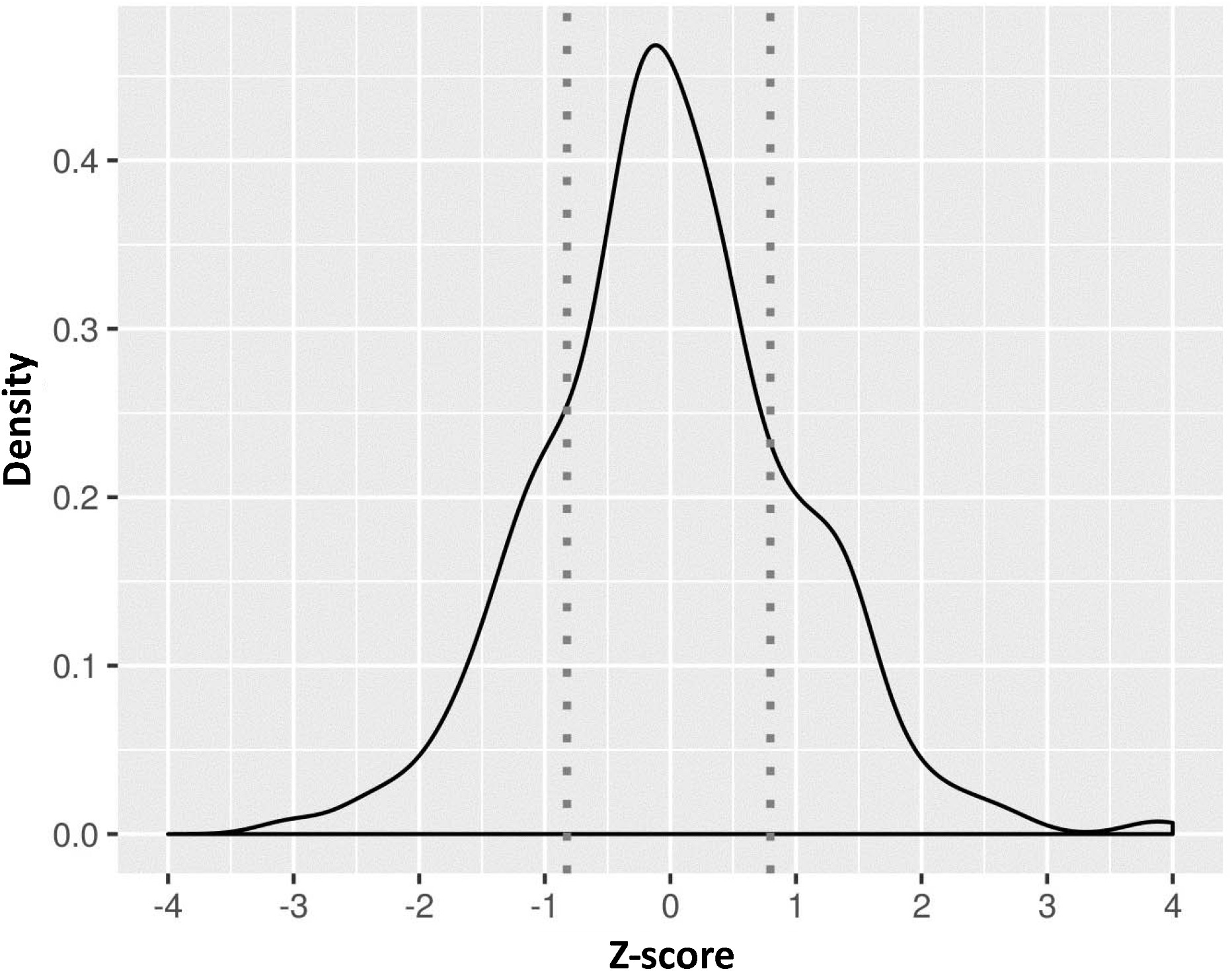
Genetic risk score distribution. Density plot for the PD genetic risk score. Z-scores reflect standard deviations from the mean. The low- and high-risk groups are indicated by dashed lines, highlighting cutoffs for the 1st and 5th quintiles, respectively.

### Survey Study

Since our IRB protocol did not permit disclosure of genome sequencing results to participants, we enrolled a largely non-overlapping sample of 231 subjects with PD from our 2 centers for a survey study, in which representative results were presented as part of clinical vignettes. Subject demographics are summarized in Table 1. A minority of subjects (n=28) completing the survey also contributed samples for genome sequencing. Prior to hearing the vignettes, the majority (81%) of subjects responded they were interested in genetic testing for PD (Figure 3). Most respondents (82%) indicated they would inform at-risk family members about the potential finding of a high-penetrance PD risk variant, such as *LRRK2*^*G2019S*^ (vignette #1). Only 2 subjects (1.3%) reported reduced interest in comprehensive genetic testing based on the potential implications for familial risk. Feedback often reflected concern for children or grandchildren.

**Figure 3.**
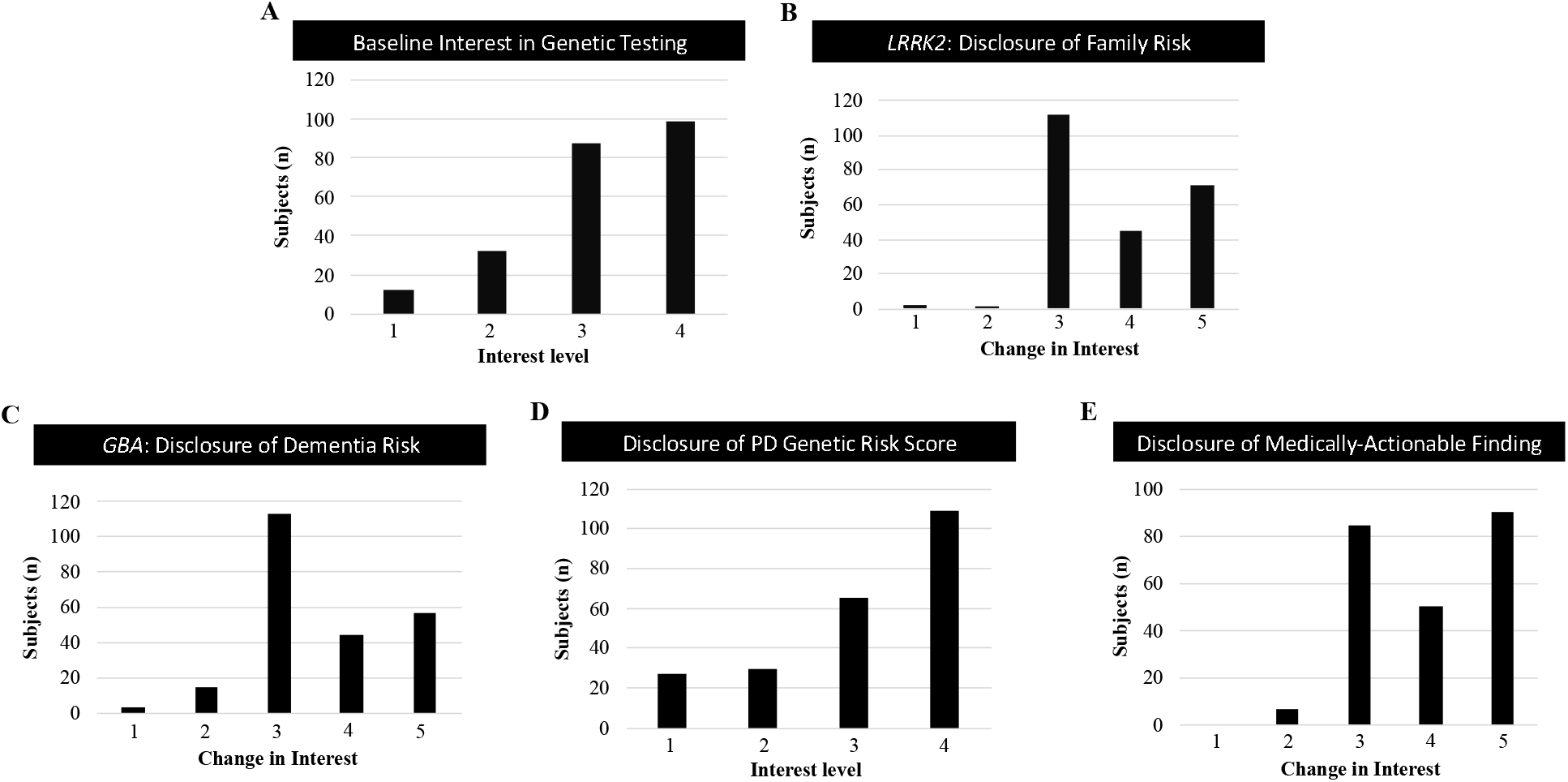
Survey Results. Results from survey questions gauging subject interest in comprehensive genetic testing at baseline (A) or following several clinical vignettes, including disclosure of a *LRRK2* variant (B, vignette #1), *GBA* variant (C, vignette #2), genetic risk score (D, vignette #3) or secondary, medically-actionable finding unrelated to PD (E, vignette #4). Most subjects reported no change or increased interest in genetic testing following disclosures. For complete survey and all results, see Supplement Information and Supplemental Table 4.

One subject commented, “I just want to know what I can do to keep my kids from getting this”, and another offered, “I don’t want to do anything that would hurt our grandchildren”. Three subjects expressed concern about potential insurance denial or job discrimination, either for themselves or their relatives.

Clinical vignette #2 focused on the potential for increased dementia risk following discovery of a *GBA* variant. Only 16 subjects (7%) reported reduced interest in genetic testing, while 101 (44%) were, in fact, more interested in genetic testing after learning that genetic variants might indicate heightened risk for PD dementia. When informed that genetic testing results, such as discovery of a *GBA* variant, would not currently change treatment decisions for the majority of patients, 75% of subjects responded that this did not change their interest in genetic testing.

For most patients, genetic testing is not expected to reveal a highly-penetrant PD risk variant. Instead, vignette #3 highlights that a genetic risk score can be computed based on a large number of genetic polymorphisms. The majority of subjects (75%) expressed an interest in learning their PD genetic risk score. Among the subjects who were not interested in their PD genetic risk score, 4 expressed concerns that PD must be predominantly influenced by environmental factors, making genetic testing less relevant. Lastly, most participants (88%) expressed a desire to receive information on medically-actionable variants unrelated to PD following vignette #4, which included discussion of a pathogenic *BRCA1* variant. In fact, 61% reported they were even more interested in genetic testing considering the possibility of learning such information.

Other general comments from subjects regarding utility of genetic testing included supporting long-term care planning, improved understanding of PD, and a desire to help discover a cure. Six subjects noted minimal interest in genetic testing due to the lack of availability of disease-modifying treatments. One volunteered, “I don’t think it should be discussed unless there is a cure”. Four subjects raised concerns about the cost of genetic testing. Six reported being unaware that PD is influenced by genetic risk factors or that genetic testing was available. Several subjects were surprised to learn that genetic risk in PD is more complicated than in other genetic disorders. One subject expressed that prior to the survey, he was concerned that genetic testing was clinically useful but was being withheld from patients due to cost. The most frequent comment at the end of the survey was a request for more information. Fifteen subjects desired more education about genetic testing during doctor visits. Seventeen requested that written material on genetics in PD be made available. Three suggested a didactic class or seminar, and 5 asked for references to websites or YouTube videos. Many expressed an increased interest in genetic testing that was specifically related to new information learned from completing the survey.

## Discussion

A substantial majority of patients with PD seen at 2 academic medical centers were interested in information about genetic risk, including the opportunity to inform at-risk family members (72%), discover their possible risk of dementia (75%), obtain their genetic risk score (75%), and learn about potential medically-actionable variants (88%). This is largely consistent with other published surveys.^21–23^ Despite rapidly expanding genetic knowledge about PD, including many established variants that affect disease risk and clinical manifestations, testing is rarely pursued in routine PD clinical care. Most commercial genetic testing panels are largely limited to monogenic causes of familial PD.^18^ Moreover, such panels often include genes with limited evidence or omit more recently discovered loci. By contrast, genome sequencing offers more comprehensive genetic analysis. As a proof of principle for clinical implementation, we obtained genome sequencing from 208 patients with PD and developed a custom gene and variant panel for filtering and analysis. In our sample, 29 subjects had reportable, diagnostic findings of either high- or moderate-penetrance PD risk or modifying alleles among 49 genes examined. These results are overall consistent with other published studies in similar PD patient cohorts.^37–39^ We also computed a 90-variant PD genetic risk score for all subjects and examined secondary findings for 59 medically-actionable genes. Importantly, compared with a targeted gene panel assay, genome sequencing coupled with focused filtering for genes relevant to PD, permits re-filtering and updated reporting based on newly published findings.^40^ For example, since completing our analyses, the ACMG has already updated its recommendations on medically-actionable findings to include 73 genes.^19^

Since sequencing was performed under a research protocol that did not allow for disclosure of genetic results to subjects, we designed a survey to assess reactions to clinical vignettes including representative findings. For the majority of subjects, we found that interest in genetic testing remained strong or increased despite potential revelations of familial PD risk, personal risk of PD dementia, or lack of immediate impact on treatment decisions. Interest in clinical genetic testing for PD is likely to increase as more patients become aware of available clinical trials of disease-modifying treatments for carriers of *LRRK2* and *GBA* variants.^41,42^ If such therapies prove successful, results of this and other larger, multi-center studies (e.g. PD GENEration^43^) will be important to inform the necessary roll-out of more routine and widespread genetic testing for PD, including the development of guidelines for effective communication with patients and families around genetics.

Based on recent studies^44^, many neurologists feel unprepared to communicate authoritatively on PD genetics, but many of the issues likely to arise in genetic counseling for PD are commonly encountered in other neurogenetic syndromes. For example, the discovery of high-penetrance *LRRK2* variants may trigger interest in “cascade testing” among family members. However, in the case of moderately penetrant variants, such as *GBA* or *APOE*, the implications for relatives may be more difficult to convey and for patients to conceptualize. Despite the potential for clinically meaningful findings, most patients are unlikely to discover responsible high- or moderate-penetrance PD risk variants, at least based on our current genetic knowledge. In our study, many subjects were surprised to learn that such negative results are the most likely outcome (86% of our sample). Inclusion of the PD genetic risk score expands the potential applicability of genetic testing, and we documented strong interest in such results. Higher genetic risk scores have been shown to be significantly associated with PD progression^45,46^, cognitive impairment^47^, and levodopa-induced dyskinesia.^48^ Nevertheless, most neurologists may not be familiar with genetic risk scores. In addition, available risk scores are derived from GWAS conducted almost exclusively in European-ancestry populations, making them poorly generalizable for other groups.

Strengths of this study include genetic sampling from 2 high-volume academic movement disorders clinic populations and inclusion of more than 200 subjects for both the genome sequencing and survey arms of the study. We also developed a custom panel for filtering and annotation of genome sequencing results based on systematic criteria and careful literature review. The clinical vignettes developed for our survey may be a valuable tool for engaging and educating patients as part of pre-test genetic counseling on potential risks and benefits. We also acknowledge some important limitations. Adoption of genome sequencing as a first-line clinical genetic test for PD may be limited by higher cost for data generation and analysis compared to single gene panels. Moreover, compared to single nucleotide variants, the detection of copy number variants may be less reliable and our analysis did not consider such alleles, which can be important contributors to PD risk^48^. For example, finding a heterozygous *PRKN* variant (R275W) in one subject, which is non-diagnostic on its own, might plausibly be in *trans* with an undetected pathogenic *PRKN* copy number variant. An additional limitation is that expert consensus annotation of PD pathogenic variants is currently lacking to guide reporting of results for many genes. Notably, in the case of *GBA* and *SMPD1*, current database annotations are focused on lysosomal storage disorders rather than PD. Lastly, it is possible that attitudes about genetic testing from our academic movement disorders clinic population may not be generalizable to community neurology practices, including those with more culturally and ethnically diverse populations. Overall, our findings highlight broad interest in comprehensive genetic testing among patients with PD and can help inform the successful integration of genome sequencing in clinical practice for PD.

## Data Availability

Complete results from the survey are available in the Supplement. For all subjects who consented for public data sharing, sequencing data are in process for release in relevant genomic databases. The complete sequencing data are also available on request by contacting the corresponding author, Dr. Shulman.

## Acknowledgments

We would like to thank all participants in this study. Kim Bambarger, Veronica Fallon, Christina Griffin and Phillip Heiser provided coordination and support for sample collection and processing. This work was supported by the Huffington Foundation, McGee Foundation, and the Jan and Dan Duncan Neurological Research Institute at Texas Children’s Hospital.

