## Supplemental Information for "Genome sequencing in the Parkinson’s disease clinic"

#### Supplemental Table 1. Parkinson's Disease Gene Panel

The panel considers 5 categories of genes/variant. Categories 1-3 are listed below, based on literature review. Category 4 consists of a published PD genetic risk score consisting of 90 single-nucleotide polymorphisms[1]. Category 5 consists of secondary, medically-actionable findings unrelated to PD, including 59 genes recommended for reporting by the ACMG[2]. PD=Parkinson's disease; YOPD=young-onset PD; LBD=Lewy body dementia; NBIA= neurodegeneration with brain iron accumulation; PSP= progressive supranuclear palsy; FAHN= fatty acid hydrolase associated neurodegeneration; PKAN= pantothenate kinase-associated neurodegeneration; BPAN= beta-propeller protein-associated neurodegeneration; FTD= frontotemporal dementia; AD=autosomal dominant; AR=autosomal recessive; XLR=X-linked recessive; XLD=X-linked dominant

| Gene | Phenotype | Inheritance | Reference |
|---|---|---|---|
| **Category 1** | | | 3 |
| *SNCA* | YOPD, LBD | AD | |
| *LRRK2* | PD | AD | |
| *VPS35* | PD | AD | |
| *PARK2* | YOPD | AR | |
| *PINK1* | YOPD | AR | |
| **Category 2** | | | 2,4 |
| *GBA* | PD, LBD | AD | |
| *SMPD1* | PD | AD | |
| *APOE* | LBD | AD | |
| **Category 3** | | | |
| *DJ-1* | PARK7, parkinsonism, psychiatric symptoms, dystonia | AR | 5–7 |
| *ATP13A2* | PARK9, Kufor-Rakeb syndrome, dystonia parkinsonism, NBIA | AR | 8–10 |
| *FBXO7* | Parkinsonism, pyramidal signs | AR | 11–13 |
| *SYNJ1* | Juvenile parkinsonism dystonia, cognitive decline, seizures | AR | 14–16 |
| *DNAJC6* | Early onset parkinsonism, intellectual disability, seizures | AR | 17–19 |
| *DNAJC13* | Parkinsonism, young or late onset, slow progression | AD | 20–22 |
| *TAF1* | Dystonia parkinsonism (DYT3, Lubag syndrome) | XLR | 23,24 |
| *SLC30A10* | Focal or generalized dystonia, parkinsonism | AR | 25–27 |
| *PLA2G6* | PARK14, dystonia parkinsonism, NBIA | AR | 28–30 |
| *GCH1* | Dopa-responsive dystonia, parkinsonism | AD | 31,32 |
| *DCTN1* | Perry syndrome, parkinsonism with central hypoventilation, PSP | AD | 33–36 |
| *VPS13C* | Parkinsonism, young onset, rapid progression | AR | 37–39 |
| *ATP1A3* | Rapid onset dystonia parkinsonism, alternating hemiplegia of childhood | AD | 40–42 |
| *PRKRA* | Dystonia-parkinsonism | AR | 43–45 |
| *GLB1* | Dystonia-parkinsonism | AR | 46–48 |
| *RAB39B* | Waisman syndrome, early onset parkinsonism, intellectual disability | XLR | 49–51 |
| *POLG* | Progressive external ophthalmoplegia, parkinsonism, ataxia, sensory neuropathy, Alper's syndrome | AR | 52–54 |
| *CSF1R* | Adult onset leukoencephalopathy, frontal lobe syndrome, parkinsonism, seizures | AD | 55,56 |
| *DNAJC12* | Hyperphenylalinemia, developmental delay, dystonia, parkinsonism | AR | 57,58 |
| *SPG11* | Spastic paraplegia, parkinsonism | AR | 59–61 |
| *TH* | Dopa-responsive dystonia | AR | 62–64 |
| *PTS* | Dopa-responsive dystonia | AD | 65,66 |
| *SPR* | Dopa-responsive dystonia | AD | 67,68 |
| *CP* | NBIA, acerulosplasminemia | AR | 69–71 |
| *FTL* | NBIA, neuroferritonopathy | AD | 72–74 |
| *FA2H* | NBIA, FAHN | AR | 75–77 |
| *PANK2* | NBIA, PKAN | AR | 78,79 |
| *COASY* | NBIA, COASY protein-associated neurodegeneration | AR | 80,81 |
| *DCAF17* | NBIA, Woodhouse-Sakati syndrome | AR | 82–84 |
| *WDR45* | NBIA, BPAN | XLD | 85–87 |
| *C19orf12* | NBIA, pallidopyramidal syndrome, dystonia, mitochondrial membrane associated neurodegeneration | AR | 88–90 |
| *PDGFB* | Familial idiopathic basal ganglia calcification | AD | 91–93 |
| *PDGFRB* | Familial idiopathic basal ganglia calcification | AD | 94,95 |
| *SLC20A2* | Familial idiopathic basal ganglia calcification | AD | 96,97 |
| *XPR1* | Familial idiopathic basal ganglia calcification | AD | 98–100 |
| *VPS13A* | Chorea-acanthocytosis | AR | 101,102 |
| *GRN* | FTD, parkinsonism | AD | 103–105 |
| *MAPT* | FTD, parkinsonism | AD | 106–108 |

#### Supplemental Table 2. Sequencing Results with Additional Information

All variants considered as pathogenic in this study were annotated in ClinVar as either "pathogenic" / "likely pathogenic" or in HGMD as "disease causing". In the case of *GBA*, database annotation is largely based on risk for Gaucher disease; however, we additionally considered 2 *GBA* variants, E365K and T408M, with strong literature support for increased risk for PD but which are non-pathogenic for Gaucher. P=pathogenic; LP=likely pathogenic; CIP=conflicting interpretations of pathogenicity; DM=disease causing.

| Gene | Transcript | Position | Variant | Subjects | ClinVar | HGMD | References |
|---|---|---|---|---|---|---|---|
| ***LRRK2*** | NM_198578.3 | chr12:40252984 | c.4937T>C:p.M1646T | 5 | CIP | DM | 109, 110 |
| | | | c.6055G>A:p.G2019S | 1 | P | DM | 110-112 |
| | | | c.1256C>T:p.A419V | 1 | P | DM | 109,113,114 |
| ***PRKN*** | NM_004562.2 | chr12:40320097 | c.823C>T:p.R275W | 1 | P | DM | 115–119 |
| ***GBA*** | NM_001005741.2 | chr1:155236376 | c.1093G>A:p.E365K | 12 | CIP | DM | 110,120,122,123 |
| | | chr1:155235252 | c.1448T>C:p.L483P | 1 | CIP | DM | 124-126,128 |
| | | chr1:155240629 | c.115+1G>A(IVS2+1) | 1 | P | DM | 127,129 |
| | | chr1:155236246 | c.1223C>T:p.T408M | 3 | CIP | DM | 110,121,123 |
| | | chr1:155235727 | c.1342G>C:p.D448H | 1 | P/LP | DM | 126,127 |
| | | chr1:155235843 | c.1226A>G:p.N409S | 2 | CIP | DM | 110,123,124,128 |
| | | chr1:155237458 | c.882T>G:p.H294Q | 1 | CIP | DM | 110,127 |

#### Supplemental Table 3. Variants of Uncertain Significance

| Gene (Transcript) | Position | Variant | Subjects (n) |
|---|---|---|---|
| ***VPS35*** (NM_018206.5) | chr16:46676573 | c.914+10T>A | 1 |
| | chr16:46679157 | c.507-1G>T | 1 |
| | chr16:46682076 | c.199+3A>G | 1 |
| | chr16:46682127 | c.151G>A:p.G51S | 1 |
| ***DNAJC13*** (NM_015268.3) | chr3:132499777 | c.4385G>A:p.R1462H | 4 |
| | chr3:132516500 | c.5560+4A>T | 22 |
| | chr3:132477972 | c.2550-9A>T | 153 |
| | chr3:132499779 | c.4387G>T:p.A1463S | 154 |
| | chr3:132450862 | c.537+15A>T | 1 |
| | chr3:132450861 | c.537+14T>A | 7 |
| | chr3:132502298 | c.4546C>T:p.R1516C | 3 |
| | chr3:132513106 | c.5385+7G>A | 3 |
| | chr3:132494190 | c.3872A>G:p.E1291G | 7 |
| | chr3:132499113 | c.4157-13T>G | 38 |
| | chr3:132505284 | c.4885-18G>A | 47 |
| | chr3:132474926 | c.2292-6G>A | 5 |
| | chr3:132478139 | c.2708G>A:p.R903K | 1 |
| | chr3:132511169 | c.5218G>C:p.E1740Q | 1 |
| | chr3:132507256 | c.5018A>G:p.Y1673C | 19 |
| | chr3:132502295 | c.4543C>T:p.P1515S | 7 |
| | chr3:132522950 | c.5796T>G:pD1932E | 1 |
| | chr3:132499231 | c.4262C>T:p.A1421V | 10 |
| | chr3:132467150 | c.2065-20A>C | 1 |
| | chr3:132505283 | c.4885-19C>T | 4 |
| | chr3:132466351 | 3:c.2021A>C:p.D674A | 2 |
| | chr3:132538225 | c.6675G>A:p.M2225I | 5 |
| | chr3:132480451 | c.2855G>A:p.R952Q | 1 |
| | chr3:132502299 | c.4547G>A:p.R1516H | 2 |
| | chr3:132514673 | c.5485+3A>C | 1 |
| | chr3:132525718 | c.6169G>T:p.A2057S | 1 |
| | chr3:132528231 | c.6424G>T:p.E2142* | 1 |
| | chr3:132466405 | c.2064+11C>T | 1 |
| | chr3:132483539 | c.3144G>T:p.L1048F | 1 |
| | chr3:132475009 | c.2369C>A:p.S790Y | 2 |
| | chr3:132491055 | c.3623+4A>G | 1 |
| | chr3:132523177 | c.5864A>C:p.D1955A | 1 |
| | chr3:132479213 | c.2710-14A>G | 1 |
| | chr3:132482333 | c.2979+3A>G | 1 |
| | chr3:132503310 | c.4813A>G:p.I1605V | 1 |
| | chr3:132456420 | c.1099+19G>A | 1 |
| | chr3:132447489 | c.294+19C>G | 1 |
| | chr3:132456401 | c.1099A>G:p.N367D | 1 |
| ***SMPD1*** (NM_000543.4) | chr11:6390606 | c.8G>A:p.R3H | 1 |
| | chr11:6390705 | c.107T>C:p.V36A | 165 |
| | chr11:6391414 | c.349G>A:p.V117M | 1 |
| | chr11:6391478 | c.413A>T:p.H138L | 1 |
| | chr11:6391523 | c.458T>A:p.L153Q | 1 |
| | chr11:6391630 | c.565A>C:p.K189Q | 70 |
| | chr11:6391631 | c.566A>C:p.K189T | 1 |
| | chr11:6391784 | c.719G>A:p.R240Q | 1 |
| | chr11:6391966 | c.901G>A:p.V301I | 1 |
| | chr11:6392060 | c.995C>A:p.P332H | 1 |
| | chr11:6393616 | c.1264-1G>T | 1 |
| | chr11:6393681 | c.1328G>A:p.R443Q | 1 |
| | chr11:6394015 | c.1460C>T:p.A487V | 6 |
| | chr11:6394233 | c.1522G>A:p.G508R | 86 |
| | chr11:6394382 | c.1671C>G:p.N557K | 1 |
| ***GBA*** (NM_001005741.2) | chr1:155235109 | c.1506-9G>A | 1 |
| | chr1:155235110 | c.1506-10T>G | 1 |
| | chr1:155236322 | c.1147G>A:p.G383S | 1 |
| | chr1:155236409 | c.1060G>C:p.D354H | 1 |
| | chr1:155237596 | c.762-18T>A | 4 |
| | chr1:155240707 | c.38A>G:p.K13R | 2 |
| ***LRRK2*** (NM_198578.3) | chr12:40235634 | c.356T>C:p.L119P | 1 |
| | chr12:40251346 | c.1073C>T:p.T358M | 1 |

| | | | |
|---|---|---|---|
| | chr12:40251455 | c.1102-10C>A | 134 |
| | chr12:40252906 | c.1182-4A>G | 1 |
| | chr12:40259538 | c.1477C>T:p.R493C | 1 |
| | chr12:40263898 | c.1653C>G:p.N551K | 31 |
| | chr12:40278187 | c.2167A>G:p.I723V | 25 |
| | chr12:40287559 | c.2689+20A>C | 1 |
| | chr12:40298479 | c.3333G>T:p.Q1111H | 1 |
| | chr12:40299212 | c.3451G>A:p.A1151T | 1 |
| | chr12:40309109 | c.4193G>A:p.R1398H | 29 |
| | chr12:40310461 | c.4348G>A:p.V1450I | 2 |
| | chr12:40313976 | c.4541G>A:p.R1514Q | 3 |
| | chr12:40314059 | c.4624C>T:p.P1542S | 11 |
| | chr12:40314114 | c.4679A>T:p.N1560I | 1 |
| | chr12:40320043 | c.4883G>C:p.R1628P | 1 |
| | chr12:40320099 | c.4939T>A:p.S1647T | 118 |
| | chr12:40320188 | c.5015+13T>A | 1 |
| | chr12:40321129 | c.5111T>C:p.F1704S | 1 |
| | chr12:40322530 | c.5509+20A>C | 1 |
| | chr12:40323151 | c.5510-9A>G | 3 |
| | chr12:40323266 | c.5616T>A:p.N1872K | 1 |
| | chr12:40340439 | c.6094T>A:p.S2032T | 1 |
| | chr12:40346884 | c.6241A>G:p.N2081D | 9 |
| | chr12:40351585 | c.6428G>A:p.R2143H | 1 |
| | chr12:40363526 | c.7153G>A:p.G2385R | 1 |
| | chr12:40364850 | c.7190T>C:p.M2397T | 185 |
| | chr12:40364879 | c.7219A>T:p.K2407* | 1 |
| | chr12:40367044 | c.7429C>T:p.R2477W | 1 |
| | chr12:40367668 | c.7487G>T:p.W2496L | 1 |
| | chr12:40367709 | c.7528C>T:p.H2510Y | 1 |
| ***PRKN*** (NM_004562.2) | chr6:162201272 | c.413-20T>C | 204 |
| | chr6:161386823 | c.1138G>C:p.V380L | 58 |
| | chr6:161360193 | c.1180G>A:p.D394N | 11 |
| | chr6:162201165 | c.500G>A:p.S167N | 12 |
| | chr6:162262692 | c.245C>A:p.A82E | 1 |
| | chr6:162054135 | c.574A>C:p.M192L | 1 |
| | chr6:161785820 | c.823C>T:p.R275W | 1 |
| | chr6:162443341 | c.140G>A:p.G47E | 1 |
| | chr6:161360169 | c.1204C>T:p.R402C | 1 |
| | chr6:162443345 | c.136G>A:p.A46T | 1 |
| | chr6:161785844 | c.799T>C:p.Y267H | 1 |
| | chr6:161569358 | c.930G>C:p.E310D | 1 |
| | chr6:161350187 | c.1310C>T:p.P437L | 1 |
| | chr6:162054183 | c.535-9T>A | 1 |
| ***DJ-1*** (NM_007262.4) | chr1:7962748 | c.-23-15T>C | 1 |
| | chr1:7970895 | c.254C>G:p.S85C | 1 |
| | chr1:7970934 | c.293G>A:p.R98Q | 3 |
| | chr1:7977638 | c.323-14A>G | 2 |
| | chr1:7985019 | c.535G>A:p.A179T | 1 |
| ***PINK1*** (NM_032409.2) | chr1:20633892 | c.344A>T:p.Q115L | 19 |
| | chr1:20638041 | c.587C>T:p.P196L | 2 |
| | chr1:20645609 | c.1009C>T:p.R337C | 1 |
| | chr1:20645615 | c.1015G>A:p.A339T | 1 |
| | chr1:20645618 | c.1018G>A:p.A340T | 20 |
| | chr1:20645700 | c.1100A>G:p.N367S | 1 |
| | chr1:20648612 | c.1231G>A:p.G411S | 1 |
| | chr1:20649169 | c.1426G>A:p.E476K | 1 |
| | chr1:20649173 | c.1430C>A:p.S477* | 1 |
| | chr1:20650447 | c.1502G>A:p.R501Q | 1 |
| | chr1:20650507 | c.1562A>C:p.N521T | 90 |
| | chr1:20650525 | c.1580T>C:p.M527T | 1 |

#### Supplemental Table 4. Survey Results

Note: Complete survey script with questions immediately follows this Table.

| Question | Responses | n (%) |
|---|---|---|
| Q1 | 1 | 98 (42%) |
| | 2 | 101 (44%) |
| | 3 | 30 (13%) |
| | 4 | 2 (1%) |
| Q3 | 1 | 12 (5%) |
| | 2 | 32 (14%) |
| | 3 | 88 (38%) |
| | 4 | 99 (43%) |
| Q4 | 1 | 190 (82%) |
| | 2 | 31 (14%) |
| | 3 | 10 (4%) |
| Q5 | 1 | 2 (1%) |
| | 2 | 1 (0.5%) |
| | 3 | 112 (48.5%) |
| | 4 | 45 (19%) |
| | 5 | 71 (31%) |
| Q6 | 1 | 3 (1%) |
| | 2 | 14 (6%) |
| | 3 | 113 (49%) |
| | 4 | 44 (19%) |
| | 5 | 57 (25%) |
| Q7 | 1 | 3 (1%) |
| | 2 | 26 (12%) |
| | 3 | 173 (75%) |
| | 4 | 16 (7%) |
| | 5 | 12 (5%) |
| Q8 | 1 | 27 (12%) |
| | 2 | 30 (13%) |
| | 3 | 65 (28%) |
| | 4 | 109 (47%) |
| Q9 | 1 | 0 (0%) |
| | 2 | 7 (3%) |
| | 3 | 84 (36%) |
| | 4 | 50 (22%) |
| | 5 | 90 (39%) |
| Q10 | 1 | 16 (7%) |
| | 2 | 204 (88%) |
| | 3 | 11 (5%) |
| Q11 | 1 | 46 (20%) |
| | 2 | 65 (28%) |
| | 3 | 44 (19%) |
| | 4 | 62 (27%) |
| | Declined | 14 (6%) |
| Q12 | 1 | 8 (3%) |
| | 2 | 67 (29%) |
| | 3 | 83 (36%) |
| | 4 | 72 (31%) |
| | 5 | 1 (<1%) |
| Q13 | 1 | 2 (1%) |
| | 2 | 3 (1%) |
| | 3 | 71 (31% |
| | 4 | 94 (41%) |
| | 5 | 61 (26%) |

### Survey: Improving Communication About Genetic Testing for Parkinson's Disease

It is possible that you have considered undergoing genetic testing for Parkinson's disease (PD). While genetic testing remains relatively uncommon during routine PD clinical care, this may change as we understand more about how genetics influences disease risk or affects the disease course in individual patients. Our goal is to learn how to better prepare PD patients for genetic testing, and how best to communicate test results. We will discuss four hypothetical "case studies" and answer questionsrelated to genetic testing in PD.

**<u>To begin, let's review some common terms related to genetic testing:</u>**

- DNA or (**d**eoxyribo**n**ucleic **a**cid) is the genetic code, which is like a blueprint for all living things.
- **Genes** are segments of DNA. Genes give our cells instructions. These instructions lead to individual traits like eye color, hair color, or in some cases, may even affect risk for diseases like PD.
- **Genetic testing** takes DNA from a blood sample and looks for changes that are associated with disease.
- **Variants** are changes that make a gene different, such as the changes that can cause disease.

1. How much do you know about genetics in PD?

| |
|---|
| 1 – nothing |
| 2 – a little |
| 3 – a moderate amount |
| 4 – a lot |

Comments:______________________________________________________________________________

2. Where did you learn what you know about genetics in PD? (please check all that apply)

☐ a doctor
☐ family and friends
☐ book
☐ school
☐ news
☐From the internet (please specify which websites): _________
☐Other (please specify): _________

3. How would you characterize your interest in having genetic testing for PD?

| |
|---|
| 1 – not interested |
| 2 – a little interested |
| 3 – moderately interested |
| 4 – very interested |

#### <u>Vignette 1</u>

Sue is 70 years old and was diagnosed with PD after noticing more difficulty getting dressed and slowed walking. Sue is worried about the risk of PD for her family since her sister recently developed a tremor. Sue has genetic testing and is found to have a *LRRK2* gene variant. Siblings and children have a 50% chance of also having a *LRRK2* variant. People with this variant have a high risk to develop PD; about 85% develop the disease by age 70.

4. If your genetic testing found a *LRRK2* variant, would you tell your family about their risk of PD?

| |
|---|
| 1 – Yes, I would tell all my family |
| 2 – Yes, but I would only tell family members with possible PD symptoms, such as tremor. |
| 3 – No, I would not tell my family members |

5. Does the possibility of discovering an increased risk of PD for your family members change your interest in genetic testing?

| |
|---|
| 1 – Much less interested |
| 2 – A little less interested |
| 3 – No change |
| 4 – A little more interested |
| 5- Much more interested |

Comments:________________________________________________

#### Vignette 2

Bob is 60 years old, and was diagnosed with PD 6 years ago. He feels his memory is "not what it used to be". He sometimes has hallucinations, but he knows they are not real. During genetic testing, Bob is found to have a glucocerebrosidase (GBA) gene variant. Individuals with *GBA* variants are 5 times more likely to develop PD. Individuals with PD who have a *GBA* variant have an increased risk of dementia and hallucinations. They may also develop PD-related disability more quickly.

6. Does the possibility of discovering an increased risk of dementia change your interest in genetic testing?

| |
|---|
| 1 – Much less interested |
| 2 – A little less interested |
| 3 – No change |
| 4 – A little more interested |
| 5- Much more interested |

Comments:________________________________________________

#### Vignette 3

Alice is diagnosed with PD at age 62. She heard about experimental research therapies for patients with GBA variants, and she therefore desires genetic testing. She is initially disappointed to learn that she does not have a GBA variant. For most patients, there is a low likelihood of discovering a high-risk gene variant, such as GBA or LRRK2, on genetic testing. In most cases, therefore, genetic testing results are unlikely to change your treatment plan.

7. Does it change your interest in genetic testing to know that your results are unlikely to change your treatment plan?

| |
|---|
| 1 – Much less interested |
| 2 – A little less interested |
| 3 – No change |
| 4 – A little more interested |
| 5- Much more interested |

Comments:________________________________________________

Although genetic testing is currently unlikely to identify single, high-risk gene variants in most patients, comprehensive testing of all genes can be used to estimate a "genetic risk score", indicating the overall risk of PD based on your genetic makeup (e.g. low, medium, or high risk).

8. How interested would you be to learn your "genetic risk score"?

| |
|---|
| 1 – not interested |
| 2 – a little interested |
| 3 – moderately interested |
| 4 – very interested |

Comments:________________________________________________

#### Vignette 4

Steve is 66 years old and has had PD for 8 years. He is worried about the risk of PD for his sisters and other family, so he undergoes comprehensive genetic testing. He is found to have no high-risk PD gene variants and overall low PD genetic risk score. However, a variant is discovered in the BRCA1 gene. This gene variant increases the risk of breast and ovarian cancer in women (as well as breast cancer and prostate cancer in men). His sisters and children have a 50% chance of also having this variant. If positive, cancer screening may be required.

9. Does it change your interest in genetic testing to know that you may discover results unrelated to PD but with significant implications for you or your family's health?

| |
|---|
| 1 – Much less interested |
| 2 – A little less interested |
| 3 – No change |
| 4 – A little more interested |
| 5- Much more interested |

Comments:________________________________________________________________________________

When having comprehensive genetic testing, you are likely to be offered the choice of only receiving results related to PD, in which case any incidental findings, such as BRCA1, would not be reported to you or your doctors.

10. If you were to have comprehensive genetic testing, what choice would you make?

| |
|---|
| 1 – Receive only results related to PD |
| 2 – Receive all results, including any findings unrelated to PD. |
| 3 – I don't want comprehensive genetic testing |

11. Finances can affect the decision to pursue comprehensive genetic testing because it can be expensive and is frequently not covered by insurance. If you are comfortable doing so, please indicate which category describes your household income in the past 12 months?

| |
|---|
| 1 – Less than $50,000 |
| 2 – $50,000-100,000 |
| 3 – $100,000-150,000 |
| 4 – More than $150,000 |

#### Feedback

12. We hope these case studies and questions have helped you better understand possible PD genetic testing results. Please tell us how much you learned from this experience

| |
|---|
| 1 – I learned nothing new |
| 2 – I learned a little |
| 3 – I learned a moderate amount |
| 4 – I learned a lot |
| 5 – I am more confused now than before |

Comments:________________________________________________________________________________

13. Overall, how has this experience changed your interest in genetic testing?

| |
|---|
| 1 – Much less interested |
| 2 – A little less interested |
| 3 – No change |
| 4 – A little more interested |
| 5- Much more interested |

14. Please tell us anything you think doctors could do better to help people understand genetic testing for PD.

---
